# Depiction of Mental Illness in Film and Association with Financial and Critical Success

**DOI:** 10.1101/2020.10.20.20215707

**Authors:** Shaan Kamal, Osama El-Gabalawy, Nathan Zhao, Jelena MacLeod

## Abstract

Film and cinema are an important part of American culture and discourse. In recent years, there have been increasing discussions around the media portrayal of suicide and psychiatric illness and its subsequent impact on prejudice towards individuals with mental health issues. To date, there is no published work quantifying the depiction of mental illness in film. In this work we use movie plot descriptions to identify movies that depict mental illness and compare their financial and critical success to all movies released during the same time period.

## Introduction

Since the Academy Awards were first held in 1927, film and cinema have been an important part of American culture and discourse. The film industry is a multi-billion dollar business, with the domestic box office total in 2019 exceeding $11 billion.^1^ For many viewers, their main exposure to overt mental illness is through characters represented in movies on television shows. For many patients struggling with their own mental health, media representation can influence their perception of themselves and of how society views them.

In recent years, media content such as the Netflix’s television series *13 Reasons Why* and Warner Bros. Pictures’ 2019 film *Joker* have ignited discourse around the media portrayal of suicide and psychiatric illness in media as well as their impact on mental health stigma and prejudice.^2,3^ There are existing guidelines for suicide portrayal in media from organizations such as the World Health Organization which exist to guide filmmakers in creating accurate portrayals of suicide while also minimizing the negative impacts such as suicide contagion of such portrayals.^4^

While there has been analysis of the stereotypes of mental illness portrayed in film, and analysis of individual films, to date there is no published work quantifying the depiction of mental illness in film at large.^5,6,7^ In this work we use the Wikipedia plot summaries of movies to identify movies that depict mental illness and compare their financial and critical success to all movies.

## Methods

The full list of movies in our analysis was scraped from BoxOfficeMojo.com. BoxOfficeMojo’s complete domestic box office data goes back to 1977, and the data was collected through August 4th, 2019, resulting in 16,333 movies included as part of our initial search^8^. In order to identify the subset of films that depict mental illness, Wikipedia plots were scraped as a proxy for the actual plot depicted in each film.The Wikipedia plot scraping was done on June 10th, 2020 and resulted in 10,491 plots being collected. Notably, 5,842 films did not have a Wikipedia plot; this was due either to the absence of a plot section or Wikipedia page in general.

To define mental illness, we used search terms derived from DSM5 diagnoses, in addition to terms such as “psychiatrist”, “suicide”, and “mentally ill” (a full list of the search terms available in the supplemental). These search terms were then identified in the scraped Wikipedia plots to classify movies as depicting mental illness.

We compared the financial and critical success of movies depicting mental illness to all movies released in the same time period. We used BoxOfficeMojo’s reported domestic total box office totals for each film as a proxy for financial success. Proxy values for critical success were calculated usingIMDB ratings and Academy Award nominations. IMDB ratings were scraped from the IMDB website on November 17th, 2019 and Academy Award information was collected from the official Academy Awards website.

## Results

Overall, 2,043 movies were identified as having plots involving mental illness out of the 10,491 overall films’ plots searched. The most common search term found was ‘suicide’ with 1,114 films plots including the term (all term counts in Figure 1, Supplemental Figure 1). The number of films depicting mental illness increased on average each year from 1977 to 2019 by 2 films per year (Figure 2). The percentage of films depicting mental illness released each year out of the total population of all films released varied each year but remained between 10-20% (Supplemental Figure 2).

**Figure 1:**
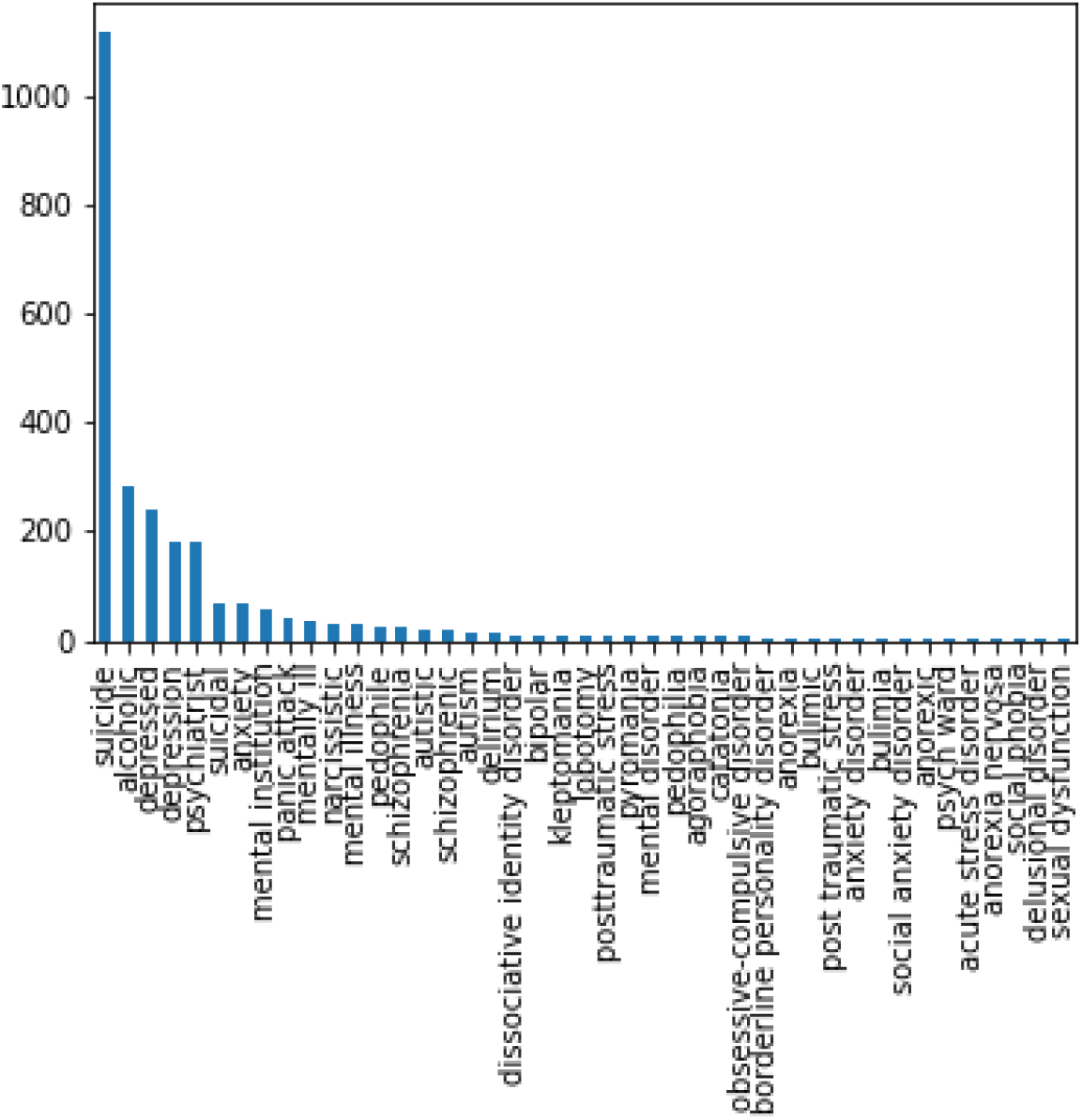
Most Commonly Found Search Terms

**Figure 2:**
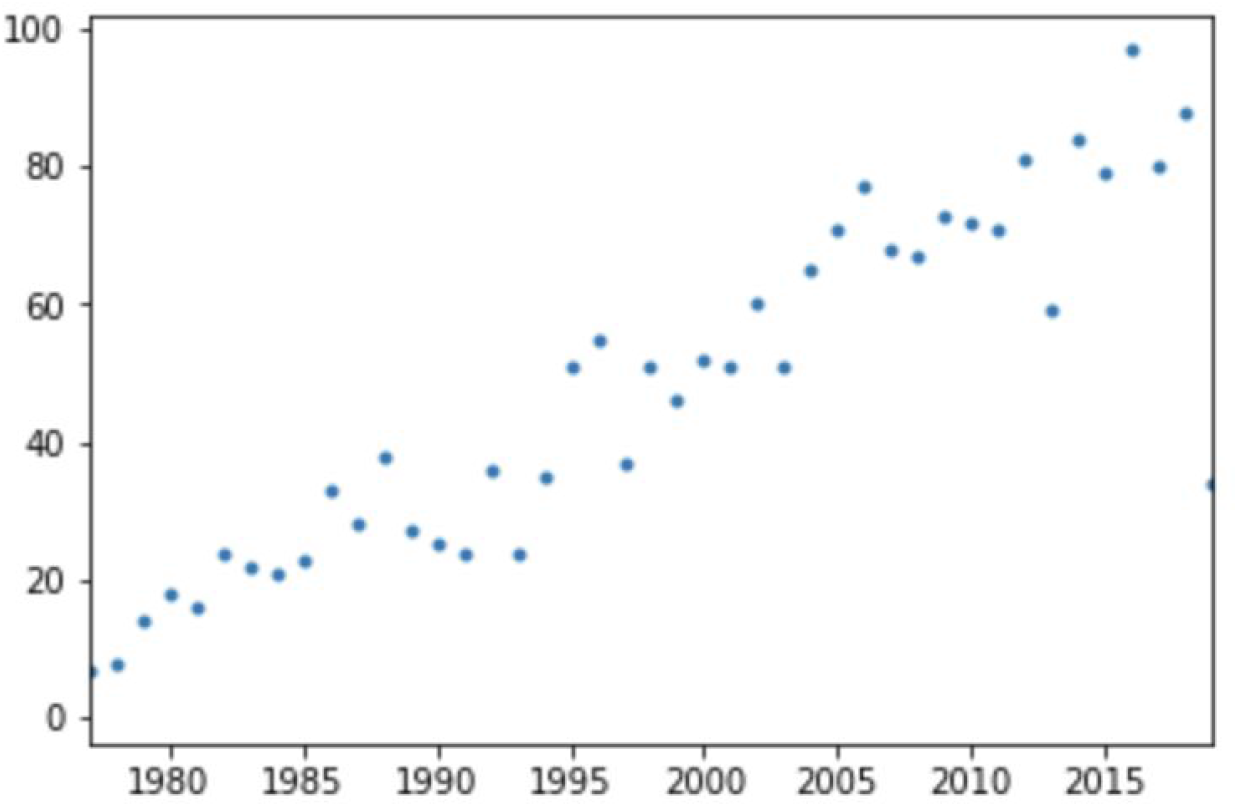
Number of Films Depicting Psychiatric Illness Released By Year

After completion of our search, we compared the financial and critical success of films depicting mental illness to all films released between 1977 and 2019. We compared the financial success of films depicting mental illness using domestic box office gross. The average gross of films depicting mental illness each year was higher than the average gross of all films in 83% of the years from 1977 to 2019 and 96.6% of films from 1990 to 2019 (Figure 3, the total domestic box office gross of each film depicting mental illness in Supplemental Figure 3). We compared the critical success of the two groups of films by using IMDB ratings and Academy Award nominations and wins as our metrics for critical success. The average IMDB rating for the films depicting mental illness released each year was higher than the average IMDB rating for all films each year over the period from 1977 to 2019 (Figure 4). The distribution of IMDB ratings for all films depicting mental illness (irrespective of year) is right shifted compared to the same distribution of all films, with the films depicting mental illness having an average rating of 6.4 and the set of all films having an average rating of 5.9 (Supplemental Figure 4, note that only 1,313 of the 10,491 films depicting mental illness had IMDB ratings that were found and included in the dataset).

**Figure 3:**
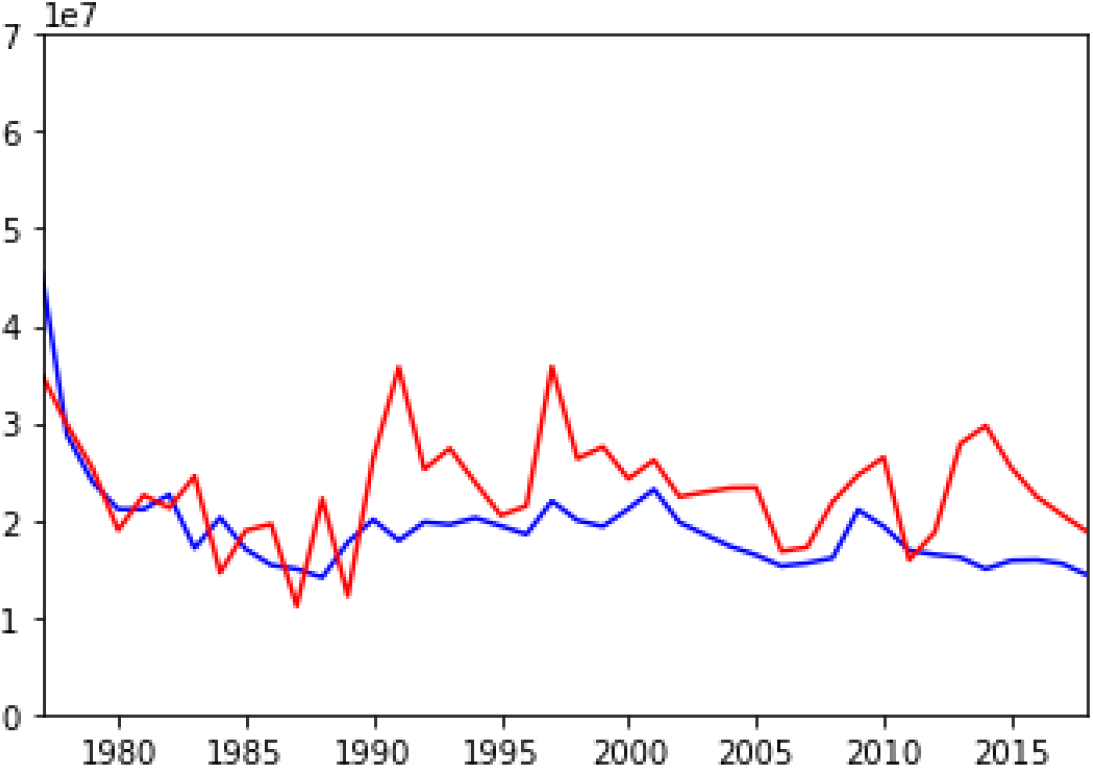
Average Box Office Gross of Films Depicting Mental Illness (red) vs All Movies (blue) by year (y-axis is 10’s of millions of dollars)

**Figure 4:**
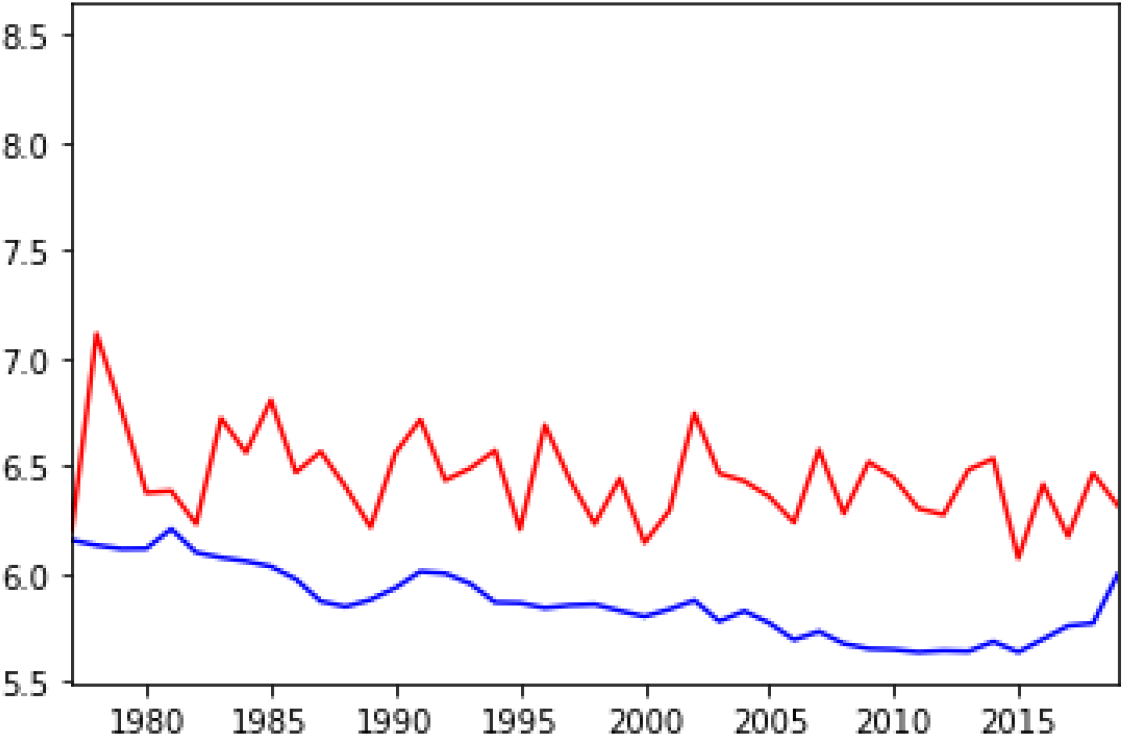
The Average IMDB rating for Films Depicting Mental Illness (red) and All Movies (blue) By Year

When looking at Academy Award nominations and wins, films depicting mental illness have consistently received nominations and wins, with the percent of nominations and wins varying each year from 1977 to 2019 (Figure 5). In total, films depicting mental illness account for 15.7% of all nominations and 17.2% of awards given out from 1977 to 2019.

**Figure 5:**
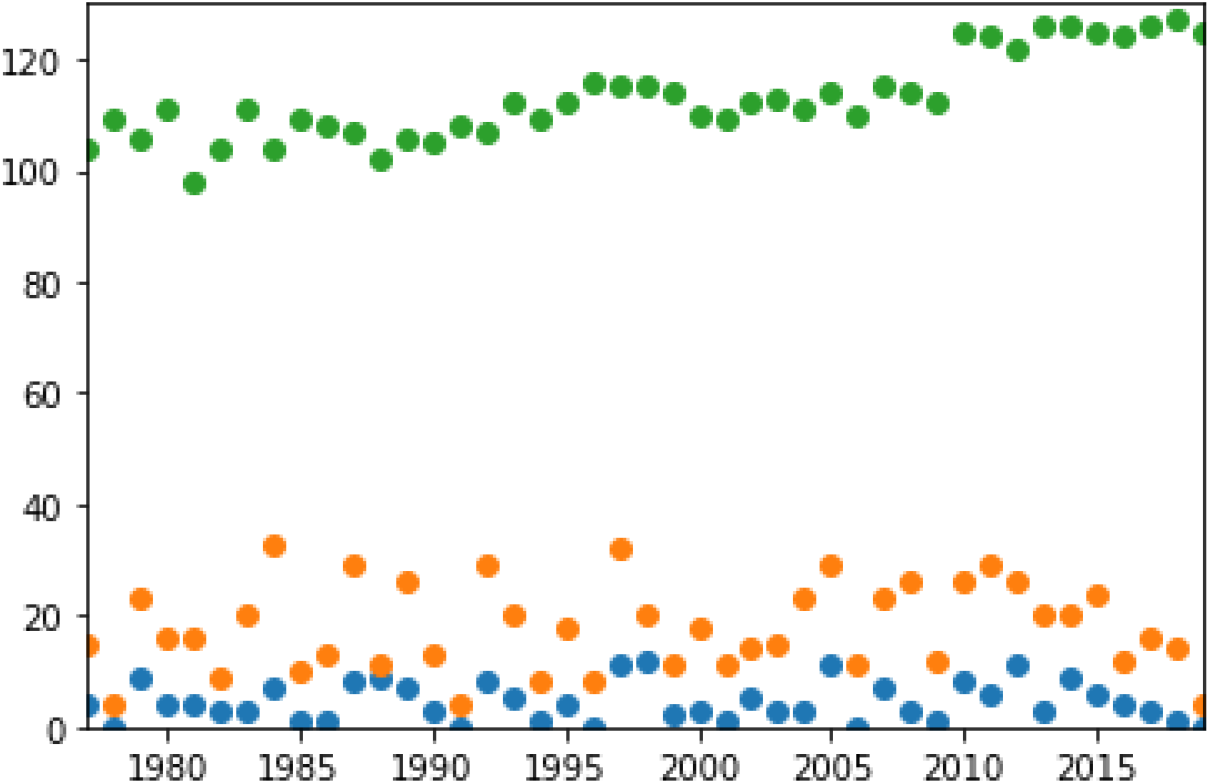
Number of Total Academy Award Nominations Given Out (Green), Nominations Given to Films Depicting Mental Illness (Orange), and Academy Awards Won by Films Depicting Mental Illness (Blue)

## Discussion

The results from this study highlight the significant depiction of mental illness in film since 1977. That films depicting mental illness have increased in number each year since 1977 and have had a higher average box than all films each year except 2011, indicating a substantial and growing interest in mental health and mental illness depictions from both the production and consumer standpoints. In addition, suicide being the most common term found further reinforces the need for widely accessible, evidence-based guidelines for accurate, sensitive depiction of suicide and mental illness in film.

The critical reception in the form of IMDB ratings for films depicting mental illness being better than the average film consistently each year and overall suggests that audiences and critics appreciate these movies overall slightly more. This is reinforced by these films also consistently receiving Academy Award nominations and wins each year despite only ∼100 such awards being given out each year and thousands of movies released each year. This again highlights that there is great interest in mental illness, but there is danger of fetishizing mental health problems or of poor depictions being celebrated.

### Limitations

Not all movies had Wikipedia plots. The plots may not have every detail as presented in the film because they are written by Wikipedia users. The box office is only domestic, not international, and does not account for the budget or marketing budget of the films.

## Conclusion

All of this information highlights how integral to popular culture and society the discussion of mental health is, how it has grown over the past decades, and how psychiatrists and mental health providers must play a role in shaping future depictions of mental illness in cinema as well as the need for evidence-based guidelines for media portrayal of all mental illness, not just for suicide.

## Data Availability

All data in the manuscript is publicly available from the sources described, our work only provides an analysis of it.

https://en.wikipedia.org/

## Author Contributions

SK and OE both collected and analyzed the data, and wrote the manuscript. All authors contributed to editing of the final manuscript.

## Supplemental

*Search Terms Used:* mental disorder, alcoholic, suicide, suicidal, psychiatrist, lobotomy, mental illness, mentally ill, mental institution, psych ward, narcissistic, Autism Spectrum Disorder, Autism, Autistic, Attention-Deficit/Hyperactivity Disorder, ADHD, AD/HD, Attention Deficit Hyperactivity Disorder, Attention Deficit Hyperactivity Disorder, Schizophrenia Spectrum, Schizophrenic, Psychotic Disorder, Schizotypal, Delusional Disorder, Brief Psychotic Disorder, Schizophreniform Disorder, Schizophrenia, Schizoaffective Disorder, Psychotic Disorder, Catatonia, Catatonic Disorder, Psychotic Disorder, Bipolar, Bipolar I, Bipolar 1, Bipolar II, Bipolar 2, Cyclothymic, Depressive Disorder, Depression, Depressed, Disruptive Mood Dysregulation Disorder, Major Depressive Disorder, Persistent Depressive Disorder, Dysthymia, Depressive Disorder, Anxiety, Anxiety Disorder, Separation Anxiety, Selective Mutism, Specific Phobia, Social Anxiety Disorder, Social Phobia, Panic Disorder, Panic Attack, Agoraphobia, Generalized Anxiety Disorder, Substance/Medication-Induced Anxiety Disorder,Obsessive-Compulsive, Obsessive-Compulsive Disorder, Obsessive Compulsive Disorder, Body Dysmorphic Disorder, Hoarding Disorder, Trichotillomania, Hair-Pulling Disorder, Excoriation, Skin-Picking, Reactive Attachment Disorder, Disinhibited Social Engagement Disorder, Posttraumatic Stress, PTSD, Post Traumatic Stress, Acute Stress Disorder, Adjustment Disorder, Dissociative Identity Disorder, DID, Dissociative Amnesia Depersonalization, Dissociative Amnesia Derealization, Somatic Symptom, Illness Anxiety Disorder, Conversion Disorder, Functional Neurological Symptom Disorder, Factitious Disorder, Pica Rumination Disorder, Pica, Avoidant Food Intake Disorder, Restrictive Food Intake Disorder, Anorexia Nervosa, Anorexia, Anorexic, Bulimia Nervosa, Bulimia, Bulimic, Binge-Eating Disorder, Sleep Terrors, Nightmare Disorder, Sexual Dysfunction, Delayed Ejaculation, Erectile Disorder, Female Orgasmic Disorder, Female Sexual Interest/Arousal Disorder, Genito-Pelvic Pain, Penetration Disorder, Male Hypoactive Sexual Desire Disorder, Gender Dysphoria, Oppositional Defiant Disorder, Intermittent Explosive Disorder, Conduct Disorder, Antisocial Personality Disorder, Pyromania, Kleptomania, Alcohol Use Disorder, Alcohol Intoxication, Alcohol Withdrawal, Phencyclidine Use Disorder, Opioid Use Disorder, Opioid Intoxication, Opioid Withdrawal, Gambling Disorder, Delirium, Paranoid Personality Disorder, Schizoid Personality Disorder, Schizotypal Personality Disorder, Antisocial Personality Disorder, Borderline Personality Disorder, Histrionic Personality Disorder, Narcissistic Personality Disorder, Avoidant Personality Disorder, Dependent Personality Disorder, Obsessive-Compulsive Personality Disorder, Voyeuristic Disorder, Exhibitionistic Disorder, Frotteuristic, Sexual Masochism Disorder, Sexual Sadism Disorder, Pedophilic Disorder, Pedophilia, Pedophile, Fetishistic Disorder, Transvestic Disorder

**Supplemental Figure 1:**
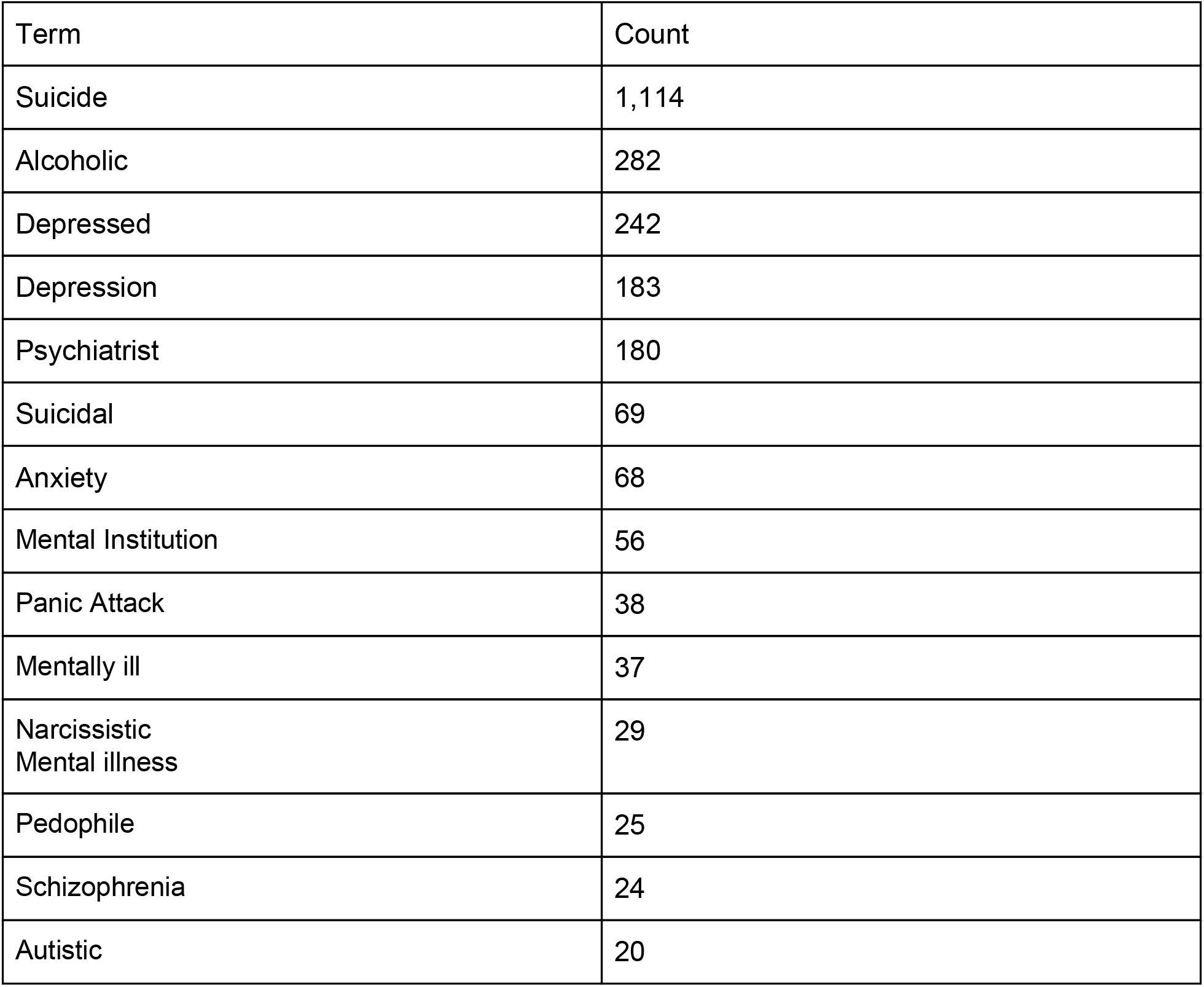
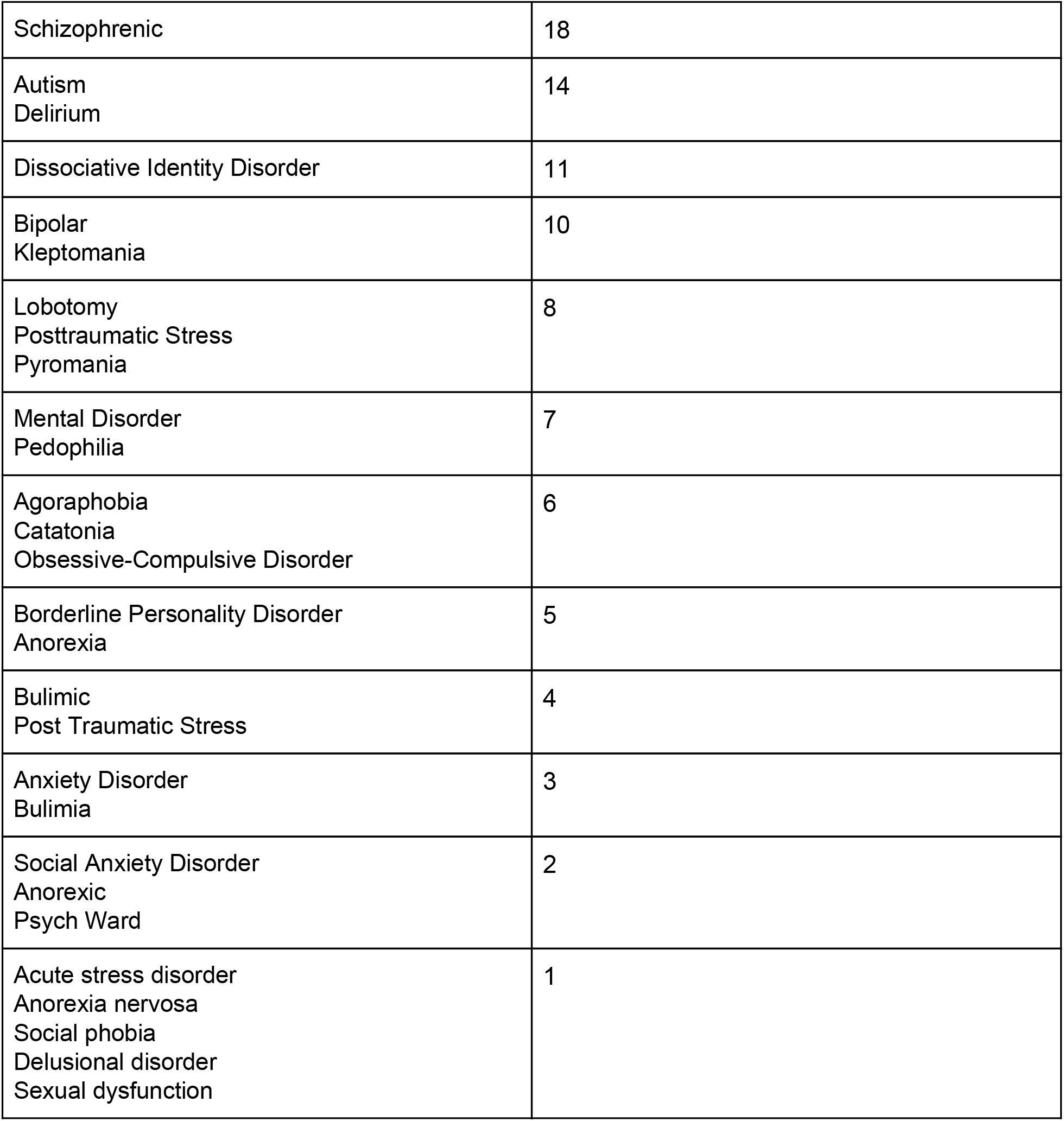
Table of Raw Data

**Supplemental Figure 2:**
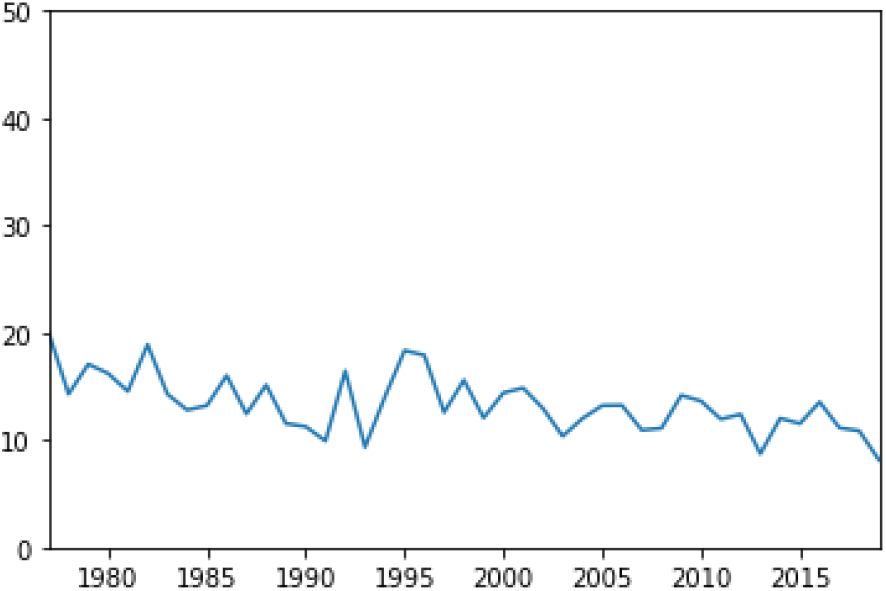
Percentage of Films Released Each year that are Films Depicting Mental Illness

**Supplemental Figure 3:**
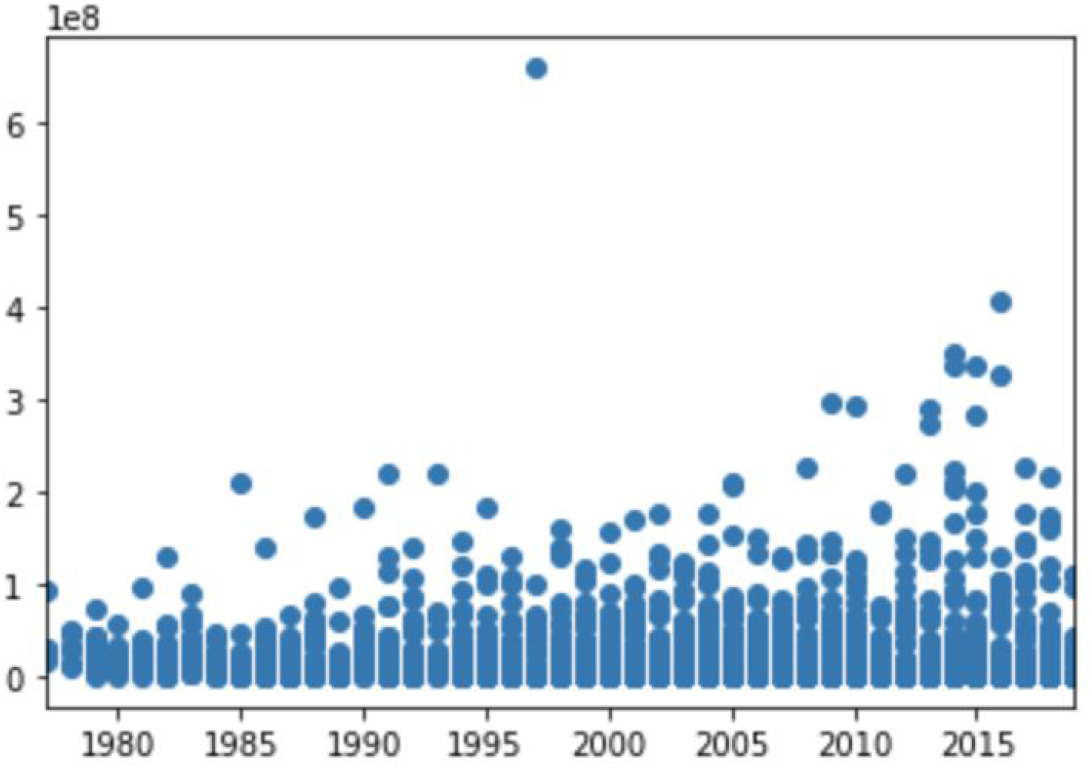
The Total Domestic Box Office Gross of Each Film Depicting Mental Illness (y-axis is 100’s of millions of dollars)

**Supplemental Figure 4:**
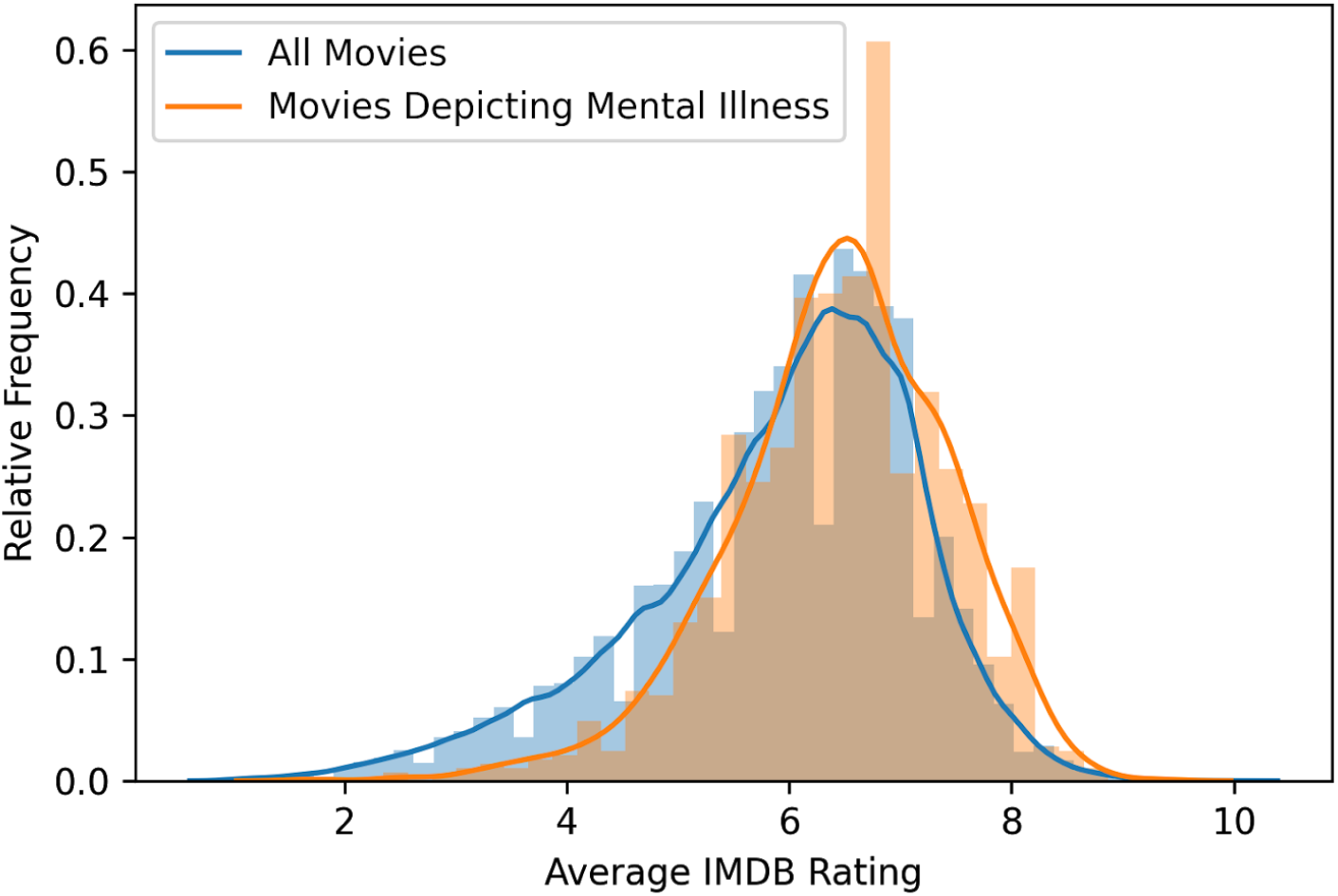
IMDB rating relative frequency distribution of movies depicting mental illness (orange, mean = 6.4, n =1313) compared to all movies (blue, mean = 5.9, n =81,273). Gaussian kernel density estimate curve overlaid for visualization.

